# What is the relationship between validated frailty scores and mortality for adults with COVID-19 in acute hospital care? A systematic review

**DOI:** 10.1101/2020.11.13.20231365

**Authors:** Theodore D. Cosco, John Best, Daniel Davis, Daniele Bryden, Suzanne Arkill, James van Oppen, Indira Riadi, Simon Conroy

## Abstract

**Background & aim:** The aim of this systematic review was to quantify the association between frailty and COVID-19 in relation to mortality in hospitalised patients.

**Methods:** Medline, Embase, Web of Science and the grey literature were searched for papers from inception to 10^th^ September 2020; the search was re-run in Medline up until the 9^th^ December 2020. Screening, data extraction and quality grading were undertaken by two reviewers. Results were summarised using descriptive statistics, including a meta-analysis of overall mortality; the relationships between frailty and COVID-19 mortality were summarised narratively.

**Results:** 2286 papers were screened resulting in 26 being included in the review. Most studies were from Europe, half from the UK, and one from Brazil; the median sample size was 242.5, median age 73.1 and 43.5% were female. 22/26 used the Clinical Frailty Scale; reported mortality ranged from 14 to 65%. Most, but not all studies showed an association between increasing frailty and a greater risk of dying. Two studies indicated a sub-additive relationship between frailty, COVID-19 and death, and two studies showed no association.

**Conclusions:** Whilst the majority of studies have shown a positive association between COVID-19 related death and increasing frailty, some studies suggested a more nuanced understanding of frailty and outcomes in COVID-19 is needed. Clinicians should exert caution in placing too much emphasis on the influence of frailty alone when discussing likely prognosis in older people with COVID-19 illness.

**Key points:**

- Frailty is being used to assess the risk of dying from COVID-19
- Emerging studies demonstrate a complex relationship between frailty and COVID-19 related deaths
- Clinicians should exert caution in placing too much emphasis on the influence of frailty in older people with COVID-19
- Researchers should ensure that frailty scales are used as designed when planning and reporting future research.

## Introduction

The COVID-19 pandemic has had a disproportionate impact upon older people. An emerging feature of the clinical response has been to use the frailty construct to estimate likely outcomes or direct treatment escalation planning [1, 2]. Frailty is a state of increased vulnerability to poor resolution of homeostasis after a stressor event, which increases the risk of adverse outcomes, including delirium, disability and death [3-5].

Where frailty has previously been studied in the critical care context, lower levels of frailty have been associated with better outcomes [6]. This data may have informed the decision by the National Institute of Clinical Excellence to encourage the use of the Clinical Frailty Scores when considering critical care escalation in older people with COVID-19 illness [2]. At the time of the NICE guidance being issued, there had been no studies validating such an approach in the context of COVID-19. Since, a number of studies have assessed outcomes from COVID-19 in older people, using various frailty scales.

The aim of this review was to synthesise emerging findings by quantifying the association between frailty and COVID-19 illness in relation to mortality in hospitalised patients.

## Methods

The full systematic review protocol has been published elsewhere (PROSPERO ID: CRD42020200445)[7]; the only change to the protocol was the extension of the search period (see below).

### Search strategies

Medline, Embase and Web of Science databases were searched with exploded MeSH headings and relevant keywords, restricted to English language. Databases were searched from inception to 10^th^ September 2020, and references were managed using Endnote software. The reference lists of included full-texts were hand-searched for additional papers. Indicative search terms are displayed below; these were modified accordingly for each database.

“Frail*”

AND

COVID-19 ((“COVID-19” OR “COVID-2019” OR “severe acute respiratory syndrome coronavirus 2” OR “severe acute respiratory syndrome coronavirus 2” OR “2019-nCoV” OR “SARS-CoV-2” OR “2019nCoV” OR (Wuhan AND coronavirus))

Grey literature was accessed by searching: Open Grey, medRxiv, bioRxiv.

A focused search was re-run in Medline on the 9^th^ December 2020 to seek out more recent studies.

### Inclusion Criteria

- Studies published from inception to 9^th^ December 2020.
- Original peer-reviewed articles, pre-prints, conference proceedings and letters to the editor reporting primary data, in any language.
- Studies reporting mortality as related to frailty in individuals diagnosed with COVID-19 in acute hospital settings.
- Frailty identified using a recognised frailty instrument.
- Participants with a positive diagnosis of COVID-19 (SARS-CoV-2 RNA-PCR positive or specialist clinical opinion).
- Participants aged 18 years or older.

### Exclusion Criteria

- Studies not involving humans.
- Articles not reporting primary data.
- Studies in which COVID-19 was self-diagnosed.

### Study Quality Assessment

Two independent reviewers (TDC and KW or SC and JvO) assessed the study quality using the Newcastle-Ottawa Quality Assessment Scale (NOS). The NOS scale uses a ‘star system’ assess the validity of studies in the domains of the selection and comparability of cohorts, and the ascertainment of either the exposure or outcome of interest. This gives rise to quality ratings:

- **Good quality:** 3 or 4 stars in selection domain AND 1 or 2 stars in comparability domain AND 2 or 3 stars in outcome/exposure domain
- **Fair quality:** 2 stars in selection domain AND 1 or 2 stars in comparability domain AND 2 or 3 stars in outcome/exposure domain
- **Poor quality:** 0 or 1 star in selection domain OR 0 stars in comparability domain OR 0 or 1 stars in outcome/exposure domain.

A maximum of 2 stars were possible in the selection domain, as cohorts were not matched for frailty.

### Data Extraction

Two reviewers (TDC and KW/IR or SC and JvO) identified and exported articles identified by the search strategy into EndNote reference software; duplicates were deleted. Independent title and abstract screens were conducted by TDC,KW or SC identifying articles for full-text extraction. Full-text screening was used to identify a final list of included studies. Relevant data were extracted by two independent researchers (JB and TDC, or SC and JvO) from the included studies into a pre-established extraction form.

### Analysis

Summary statistics for age were combined, after converting medians/IQRs into means and standard deviations using Wan’s method [8]. Overall mortality was summarised using meta-analysis, with heterogeneity assessed using the I-squared statistic [9]. A meta-analysis summarising the effect of frailty on COVID-19 mortality was planned but the heterogeneity in study designs, frailty tools used and reporting of mortality made this impossible.

### Ethics and funding

No ethical approval was required for this work.

Daniel Davis is funded through a Wellcome Intermediate Clinical Fellowship (WT107467).

Theodore D Cosco is funded through a Michael Smith Foundation for Health Research Scholar Award (SCH-2020-0490).

James van Oppen is funded through a National Institute for Health Research Doctoral Research Fellowship (NIHR300901).

## Results

The initial searches identified 2276 records of which 650 were duplicates, leaving 1626 papers for review. After scrutinising the titles and abstracts against the eligibility criteria, 36 papers were retained for full-text review; a further 10 papers were identified on re-running the search in Medline, leading to 26 papers being included for data abstraction (Figure 1).

**Figure 1.**
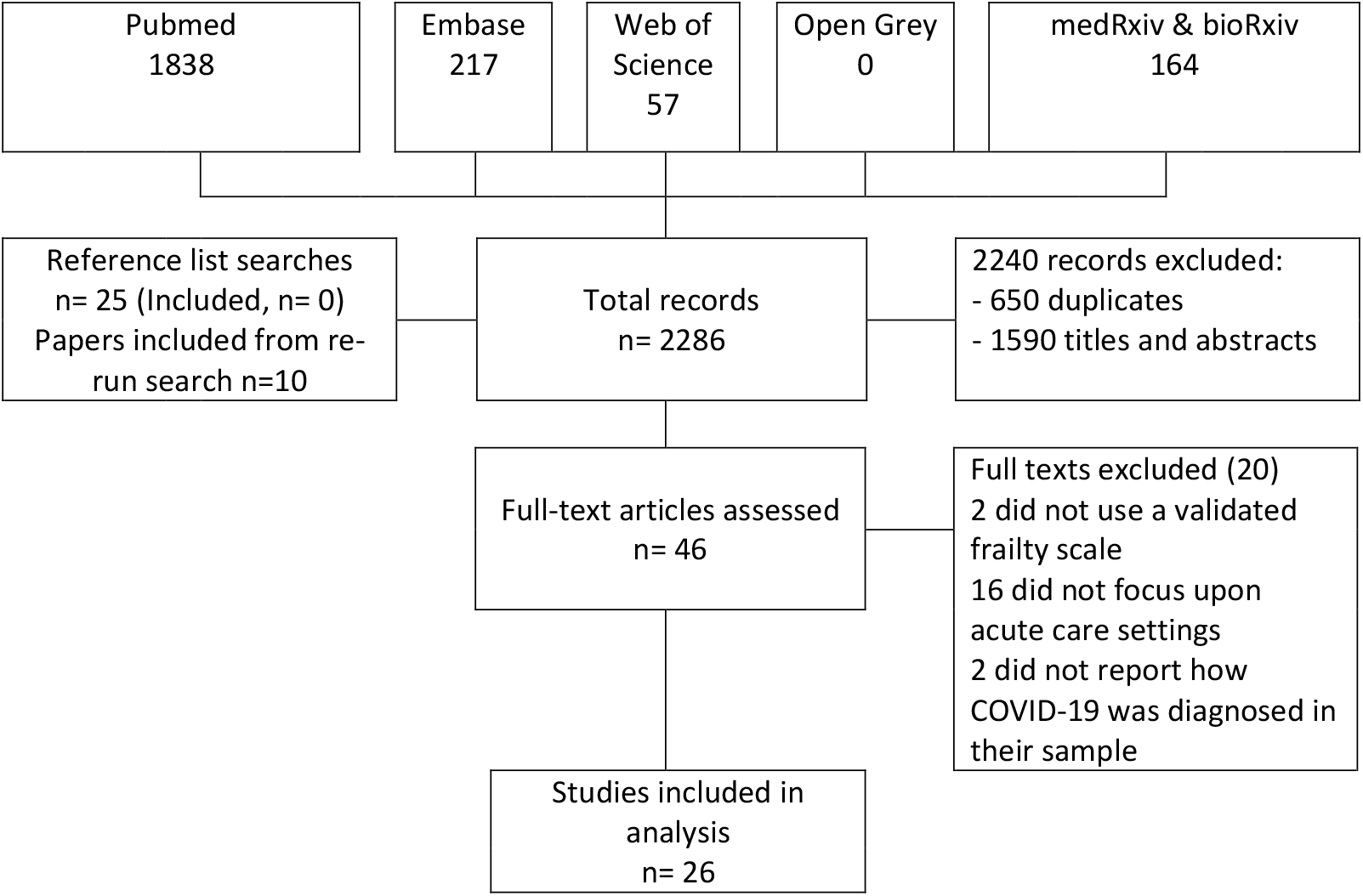
Study selection

The summary characteristics are shown in Table 1.

**Table 1.** Summary characteristics of retained studies examining frailty and COVID-19 related outcomes

| Author | Country | Setting | Sample size | Age; mean (SD) | Proportion female | Proportion White | Frailty measure | COVID diagnosis | NOS grading |
| --- | --- | --- | --- | --- | --- | --- | --- | --- | --- |
| Apea [16] | UK | Five acute hospitals | 1996 | 62.2 (17.4) | 39% | 35% | CFS | PCR | 6 |
| Aw [22] | UK | Acute hospital | 677 | 81.1 (8.1) | 46% | 81% | CFS | PCR | 6 |
| Baker [23] | UK | Acute hospital | 316 | 72.7 (17.1) | 45% | 96% | CFS | PCR | 6 |
| Brill [24] | UK | Acute hospital | 410 | 81.1 (8.1) | 65% | 60% | CFS | PCR | 4 |
| Chinnadurai [25] | UK | Acute hospital | 215 | 72.0 (16.4) | 38% | 87% | CFS | PCR | 5 |
| Cobos-Siles [26] | Spain | Acute hospital | 656 | 82.7 (10.5) | 43% | Not stated | CFS | PCR | 6 |
| Conway [27] | UK | Acute hospital | 71 | 70.7 (16.6) | 42% | 99% | CFS | PCR | 4 |
| Crespo [10] | Spain | Renal transplant cohort, acute hospital | 16 | 59.7 (12.6) | 6% | Not stated | Fried | PCR | 4 |
| Davis [28] | UK | Acute hospital | 222 | 82 (range 56–99) | 67% | Not stated | CFS | PCR | 5 |
| De Smet [29] | Belgium | General hospital | 81 | 70.3 (20.1) | 59% | Not stated | CFS | PCR | 6 |
| Doglietto [11] | Italy | Patients with COVID undergoing surgery | 41 | 82.7 (10.5) | 56% | Not stated | CFS | PCR | 4 |
| Frost [30] | UK | Seven acute hospitals | 749 | 85.3 (6.8) | 32% | Not stated | CFS | PCR | 6 |
| Hagg [18] | Sweden | Acute hospital | 250 | 81.0 (8.6) | 52% | Not stated | CFS/HFRS | PCR/clinical | 7 |
| Hewitt [19] | Italy/UK | 11 acute hospitals (10 England, 1 Italy) | 1564 | 76.0 (5.2) | 42% | Not stated | CFS | PCR/clinical | 6 |
| Hoek [31] | Netherlands | Multi-centre - solid organ transplant recipients | 23 | 60.7 (15.0) | 22% | 61% | CFS | PCR | 4 |
| Knights [32] | UK | General hospital | 108 | 69.3 (16.3) | 39% | 76% | CFS | PCR | 47 |
| Kundi [15] | Turkey | All acute hospitals in Turkey | 18234 | 74.1 (7.4) | 53.4% | Not stated | HFRS | PCR | 7 |
| Marengoni [12] | Italy | COVID-19 special hospital | 165 | 69.3 (14.5) | 39% | Not stated | CFS | PCR/clinical | 7 |
| Mendes [13] | Switzerland | COVID-19 special hospital | 235 | 86.3 (6.5) | 57% | 100% | CFS | PCR/clinical | 6 |
| Miles [33] | UK | Acute hospital | 217 | 59 | 38% | Not stated | CFS | PCR | 6 |
| Owen [20] | UK | Acute hospital | 301 | 68.7 (15.6) | 44% | Not stated | CFS | PCR/clinical | 6 |
| Poco [14] | Brazil | COVID-19 special hospital | 711 | 66 (-) | 43% | Not stated | CFS | PCR/clinical | 6 |
| Rawle [34] | UK | Acute hospital | 134 | 80.0 (6.8) | 46% | 76% | CFS | PCR | 4 |
| Steinmeyer [21] | France | Acute hospital | 94 | 85.5 (7.5) | 55% | Not stated | Frail<br>Non-Disabled<br>survey<br>(FIND) | PCR | 5 |
| Tehrani [35] | Sweden | Acute hospital | 255 | 66 (17) | 41% | Not stated | CFS (7<br>point<br>scale) | PCR | 7 |
| Thompson [36] | UK | Acute hospital | 470 | 78.8 (8.3) | 46% | 83% | CFS | PCR | 6 |

Thirteen of the 26 studies were from the UK, 13 from other European countries and one from Brazil. All studies reported findings from acute hospitals (secondary care), with Crespo *et al* [10] reporting specifically on renal transplant recipients, Doglietto on surgical patients [11] and three studies reported from COVID-19 dedicated hospitals (Marengoni [12], Mendes [13], Poco [14]) ; all the other studies reported outcomes for general, acute medical care. All studies described outcomes in people with clinically diagnosed and PCR-confirmed COVID-19, with the exception of Miles (contemporaneous matched controls), Owen and Aw (clinical and PCR positive versus clinically positive only) and Doglietto (historical matched controls). Most of the studies was fair-good on the Newcastle-Ottawa Quality Assessment Scale (Table 1).

The median sample size was 242.5 (IQR 108-656); the largest study reported on over 18,000 participants from Turkey (Kundi[15, 16]). Overall, the median age of included participants was 73.1 years (IQR 69.3-81.1) and 43.5% were female. Where reported, the majority of studies reflected white participants, except for Apea [16] which had a majority of non-white participants. Frailty was assessed using the Clinical Frailty Score (CFS) in 22 studies, one used the Hospital Frailty Risk Score (HFRS), one used both CFS and HFRS, one Fried’s frailty phenotype and one used the Frail Non-Disabled survey. Infection causing COVID-19 illness was confirmed using clinical features and a positive PCR in all studies though Hagg [18], Hewitt [19], Marengoni [12], Mendes [13], Owen [20], Poco [14] and Steinmeyer [21] also included people with clinical diagnoses but negative PCR tests.

Mortality was varied widely across the studies, ranging from 14 to 65%; studies reported mortality at different time points (5-60 days), some reported in-hospital deaths only and others all deaths in and outside of the hospital over the follow period. A descriptive summary is shown in Table 2 and a meta-analysis summarising overall mortality in Figure 2 (random effects was used as heterogeneity was high, I-squared 97.3%, p=0.000).

**Table 2.** Descriptions of mortality outcomes

| Author | Frailty measure used | Overall cohort mortality | Follow up (days unless otherwise stated) | Associations of frailty with mortality |
| --- | --- | --- | --- | --- |
| Mortality reported using hazard ratios (95% CI) |  |  |  |  |
| Apea [16] | CFS | 28.7% | 30 | Covariates in adjusted analysis: age, sex, ethnicity, smoking, BMI, and IMD<br>CFS 1-2: reference category<br>CF 3-4: 1.61 (0.82-3.16)<br>CFS 5-6: 1.84 (0.93-3.64)<br>CSF 8-9: 3.25 (1.49-7.06) |
| Aw [22] | CFS | 40.0% | 34 | Covariates in adjusted analysis: age, sex, ethnicity, IMD, previous hospital admissions in 2019 and NEWS-2<br>CFS 1-3: reference category<br>CFS 4 1.30 (0.76–2.21)<br>CFS 5 1.19 (0.70–2.03)<br>CFS 6 2.13 (1.34–3.38)<br>CFS 7–9 1.79 (1.12–2.88)<br>Sensitivity analyses: association between frailty and mortality was similar when cases were confined to RT-PCR positive cases. |
| Chinnadurai [25] | CFS | 40.0% | 5 (IQR 2–10) | Comparing CFS ≥5 to CFS <5, the unadjusted hazard ratio for death was 3.45; 95%CI: 1.76 - 6.79; p < 0.001) |
| Hagg [18] | CFS/HFRS | 24% | 25 | Covariates in adjusted analysis: age, sex, comorbidities, HFRS and acute kidney injury<br>CFS ≥5 vs. CFS<5 hazard ratio 1.85 (0.97–3.52)<br>HFRS hazard ratio 1.00 (0.91–1.10), adjusted for age and sex.<br>No modelling undertaken on non-COVID controls |
| Hewitt [19] | CFS | 27.2% | 28 | Covariates in adjusted analysis: age, sex, smoking, C-reactive protein, diabetes, coronary artery disease, hypertension, renal function<br>CFS 1-2: reference category<br>CFS 3-4: 1.55 (1.00–2.41)<br>CFS 5-6: 1.83 (1.15–2.91) |
|  |  |  |  | CFS 7–9: 2.39 (1.50–3.81) |
| Marengoni [12] | CFS | 25.6% | To death or discharge.<br>Max: 40 | Covariates in adjusted analysis: age, sex, primary education, number of chronic diseases<br>For each 1 point increase in the CFS score, the hazard ratio for death was 1.30 (1.05-1.62) |
| Mendes [13] | CFS | 32.3% | To death or discharge:<br>Mean 12.8<br>(SD 7.6) | Covariates in adjusted analysis: sex<br>For each 1 point increase in the CFS score, the hazard ratio for death was 1.46 (1.24-1.70)<br>Comparing CFS ≥5 to CFS <5, adjusted for sex, the hazard ratio for death was 4.39 (1.60-12.02) |
| Miles [33] | CFS | 51.2% | 60 | Covariates used in the adjusted analysis included age, sex, ethnicity, IMD<br>For each 1 point increase in the CFS score, the hazard ratio for death was 1.88 (1.37-2.59)<br>The different associations with frailty according to COVID-19 status was confirmed by demonstrating an interaction term (HR 0.51, 95% CI 0.37 to 0.71) |
| Owen [20] | CFS | 42.9% | 30 | Covariates in adjusted analysis: age, sex, acuity and comorbidities. Compares results in those with PCR confirmed COVID-19 only.<br>CFS 1-3: reference<br>CFS 4-5: 2.12 (0.86-5.18)<br>CFS 6: 1.69 (0.67- 4.28)<br>CFS 7-8: 2.36 (0.96- 5.76)<br>CFS 9: 11.97 (3.70- 38.72)<br>CFS Not Recorded 2.14 (0.89-5.13)<br>In COVID-19 positive individuals, the interaction between COVID-19 status and CFS suggests a sub-additive relationship. |
| Mortality reported using odds ratios |  |  |  |  |
| Cobos-Siles [26] | CFS | 19.5% | 33 | Comparing mild to very severely frail older people, the odds ratio for death was 8.73 (95% CI 1.37–55.46) |
| De Smet [29] | CFS | 23.5% | 48 | Covariates included in adjusted analysis: age, LDH, RT-PCR<br>For each 1-point increase in CFS, the odds of being dead at follow up increased by 1.75 (5% CI 1.1-3.4) |
| Kundi [15] | HFRS | 18.2% | In-hospital mortality – | Covariates included in the adjusted analysis: age, sex and comorbidities |
|  |  |  | maximum follow-up possible 103 days | For each 1 point increase in the HFRS, in-hospital mortality risk increased by an odds ratio of 1.036 (1.029-1.043)<br>By HFRS categories, the adjusted risk of in-hospital mortality compared to low HFRS category (HFRS<5) was: intermediate (HFRS 5-15) 1.482 (1.334-1.646); high (HFRS >15) 2.084 (1.799-2.413) |
| Rawle [34] | CFS | 64.9% |  | The risk of death was associated with an odds ratio of 2.68 (96% CI 1.26,-6.49) for each 1 point increase in CFS. |
| Tehrani [35] | CFS (7 point scale) | 27.5% | 60 | Covariates used in the adjusted analysis included age, and chronic kidney disease<br>For each 1 point increase in the 7 point CFS scale, the odds ratio for death was 2.07 (1.47–2.92) |
| Thompson [36] | CFS | 36.0% | 30 | Median CFS was significantly higher in non-survivors (6 IQR 4-7 vs. 3 IQR 2-5 for survivors. In the multivariate analysis adjusting for age, hypertension, cancer, CRP, platelet count, acute kidney injury and >50% total lung field infiltrates, frailty was not a significant predictor. |
| Other descriptions of mortality as an outcome |  |  |  |  |
| Baker [23] | CFS | 25.6% | 28 | Patients who died without ventilatory support had a median (IQR) CFS score of 7 (6 – 7). |
| Brill [24] | CFS | 42.2% | 28 | People aged 80+ that died were more frail (median (IQR) CFS 6 (5, 7) vs. 5 (4, 6), p= 0.002 |
| Conway [27] | CFS | 14.1% | Maximum 14 days | Patients who died had higher CFS scores (5.75 v 3.36, p=0.005) |
| Crespo [10] | Fried | 50.0% | 14 | Mortality if Fried >0 was 5/7 (62.5%) |
| Davis [28] | CFS | 42.8% | 30 | Frailty was associated with a higher chance of dying (46.1% mortality when CFS≥5 vs 32.7 when CFS≥4, p=0.02) |
| Doglietto [11] | CFS | 19.5% |  | No data on CFS associated mortality (used as a case-mix adjuster) |
| Frost [30] | CFS | 40.1% | 30 | Univariate difference in CFS score (median and IQR):<br>at 72-hours: 3 (2-6) alive versus 6 (4-7) deceased<br>at 30-days: 3(2-5) versus alive 5 (3-6) deceased |
| Hoek [31] | CFS | 21.7% |  | Mean CFS was 5.8 in those that died |
| Knights [32] | CFS | 31.5% | 30 | Median CFS was higher in patients over 65 who died (5, IQR 4–6) than in survivors (3.5, IQR 2–5) p<0.01). |
| Poco [14] | CFS | 37% | Median length of stay 11 | Mortality among older adults CFS≥5 = 58% vs. CFS<5 = 42%; P=0.003 |
| Steinmeyer [21] | Frail Non-Disabled survey (FIND) | 18.1% | Mean length of stay 12.0 (SD 5.5; range 2-31) | In a univariate Cox regression, frailty was not associated with mortality (25% mortality among 'robust' individuals vs 15% mortality among 'frail' and 'dependant'. |

**Figure 2.**
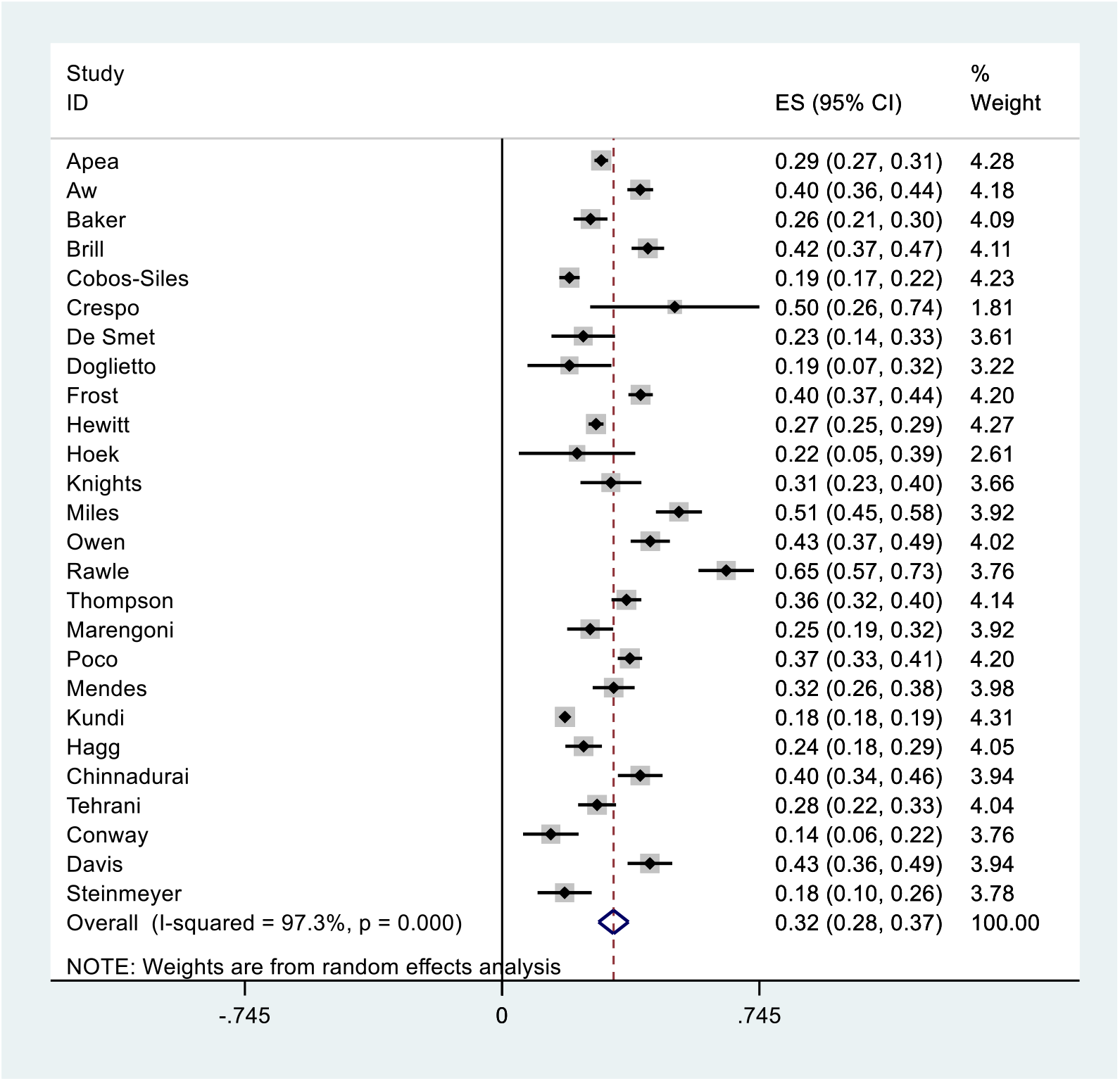
Random effects meta-analysis showing overall mortality (note high heterogeneity)

Nine studies reported mortality over time using hazard ratios to describe the effect of frailty (Apea, Aw, Chinnadurai, Hagg, Hewitt, Marengoni, Mendes, Miles, Owen); all used the CFS and with the exception of Chinnadurai, adjusted for important baseline covariates. However, these studies used different cut-points of the CFS, as well as different covariates, which meant combining the results in a meta-analysis was not possible. These studies did however all show an increase in the risk of dying with increasing levels of frailty, although Miles and Owen both found a ‘sub-additive’ effect in the most severe frailty groups, in which the increased risk of dying was less than might have been expected.

Six studies (Cobos-Siles, De Smet, Kundi, Rawle, Tehrani, Thompson) reported mortality risk as odds ratios, some using the CFS others the HFRS, again with a range of covariates and different CFS categories. It was not possible to combine these data, however with the exception of Thompson *et al*, they all showed an increased odds of dying with increasing levels of frailty.

The remaining 11 studies used a range of frailty measures in different ways to describe some aspect of COVID-19 related mortality, such that combining results would not be clinically meaningful. Most of these studies with the exception of Steinmeyer *et al* tended reported an associations between increased risk of dying and increase levels of frailty.

In summary, the majority of studies found a positive association between increasing frailty and COVID-19 related mortality – but not all. Miles and Owen both found an interaction between frailty and PCR testing that attenuated the expected mortality associated with increasing frailty. Steinmeyer *et al* and Thompson *et al* found that frailty was not a significant predictor in an adjusted analysis.

## Discussion

### Summary

This systematic review identified 26 studies assessing the influence of frailty on COVID-19 related mortality in hospitalised patients. The overall quality of the studies was reasonable, and the majority of studies showed that in older people hospitalised with COVID-19 illness that frailty was associated with COVID-19 related mortality. However, this was not consistent across all cohorts, with some showing a more complex interaction between frailty and COVID-19 status: two studies found a sub-additive interaction with frailty)i.e. that the mortality seen in severely frail older people was not as high as expected and that excess mortality was observed in those relatively fitter). This may relate to a selection effect, as policy and practice during the pandemic emphasised avoiding hospitalisation in many settings (e.g. national lockdowns). Patients with higher frailty scores are more likely to represent care-home residents, in whom COVID-19 illness might be managed in the community [37]. Treatment effects varied over time, for example, greater or lesser use of critical care or treatment escalation plans, or the introduction of ‘new’ treatments such as Dexamethasone, which could have affected outcomes. Less frail patients may have had more aggressive treatment than those with increased levels of frailty and this practice may have changed over time and varied between centres. Taken together, whilst the bulk of the studies find the ‘expected’ relationship between frailty and COVID-19 mortality, our findings suggest a more nuanced understanding of frailty and outcomes in COVID-19 is needed.

### Strengths and weakness

This review was methodologically robust according to the Quality of Reporting of Meta-analyses (QUOROM) and PRISMA reporting guidelines. It is possible that in this new field, emerging studies not yet published may have been missed, although we searched pre-print collections in an effort to minimise this risk, as well as updating the search in December 2020. The British Geriatrics Society has agreed to host a live update of this review so that future studies can be incorporated into the analysis [INSERT www once available]. Whilst the individual papers included in the review were of fair-good quality, frailty (its operationalisation and reported cut-points) and mortality were reported variably across the studies, making meta-analysis and comparisons difficult.

Most of the studies were from Europe - mostly the UK - which may limit generalisability to other health systems. We focused upon studies reporting outcomes for hospitalised patients, so we cannot make any comment about COVID-19 related risk in the wider population, in particular in care homes or population samples.

We did not examine other risk scores designed to predict outcomes from COVID-19, such as those looking at comorbidities or biomarkers [30, 38-40], as these are separate constructs from frailty. In clinical practice, both physiological risk scores and frailty risk scores would be used together to inform prognostication, and future work might compare the relative merits of combined risk scoring.

We focused upon mortality, but outcomes such as function, cognition or quality of life are equally, if not more important, especially for older people [41]. However, in this relatively early stage of the COVID-19 pandemic, we anticipated that there would be very few studies reporting such outcomes, though this will be an important area upon which to focus in the future.

### Relationship to existing literature

The CFS appears to perform similarly to other predictors of mortality in the context of COVID-19, such as the Palliative Performance Scale [39], but perhaps less well than the 4C Mortality Score, developed and validated specifically in COVID-19 cohorts [40].

Whilst mortality in hospital may be related to frailty, wider determinants of health have an important impact upon country specific survival rates. Paradoxically, 1% decrease in pre-existing all-cause mortality is associated with a 4.1% increase in the COVID-19 death rate in those ≥60 years of age, thought to be related to an unhealthy survivor effect i.e. longevity at the price of dependency and increased susceptibility to COVID-19 (e.g. care home populations) [42].

### Implications for research

Larger, more robust studies examining the relationship between COVID-19 and frailty are needed to resolve the limitations of the existing papers. Future studies should preserve the integrity of frailty scales so that comparisons can be made across studies [43], and should take account of the apparent interaction between frailty and COVID-19 testing [20, 33].

### Implications for clinical practice

Clinicians should exert caution in placing too much emphasis on the influence of frailty alone when discussing likely prognosis in older people with COVID-19 infection. No tool should be used in isolation to direct clinical care, though frailty scores can form part of a more holistic assessment to inform a shared decision making discussion. Frailty can be useful in identifying the risk of complications such as delirium - increasingly being recognised as a high risk scenario [21, 44, 45] – and further frailty or deconditioning [46]. Updated clinical guidance on frailty and COVID, as well as other resources are available here: https://www.criticalcarenice.org.uk/ and the British Geriatrics Society will maintain a live web-repository of COVID and frailty studies [HERE].

## Data Availability

All of the primary studies are already available in the public domain and referenced in the manuscript.

## Appendix 1 Grading of papers using the NOS scale

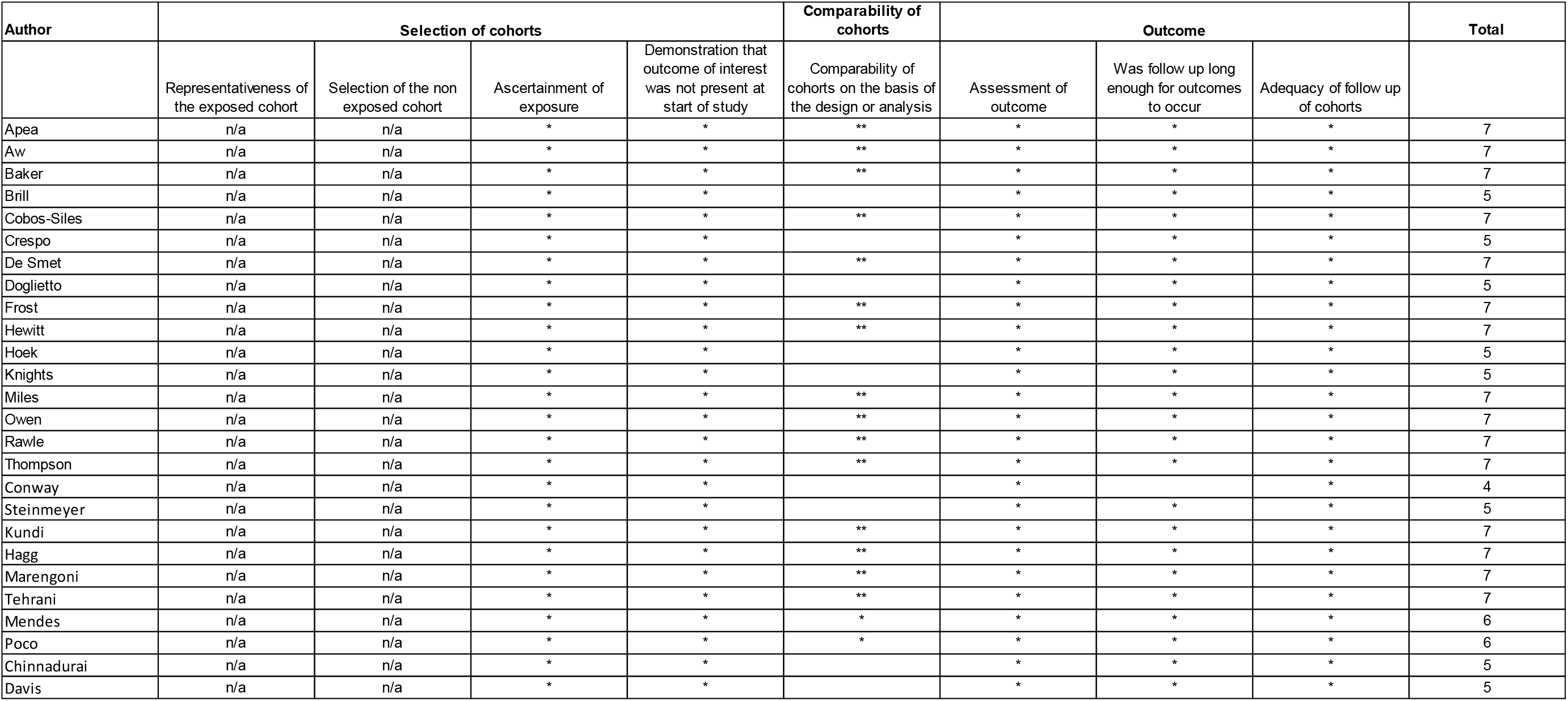

| Author | Selection of cohorts |  |  |  | Comparability of cohorts | Outcome |  |  | Total |
| --- | --- | --- | --- | --- | --- | --- | --- | --- | --- |
|  | Representativeness of the exposed cohort | Selection of the non exposed cohort | Ascertainment of exposure | Demonstration that outcome of interest was not present at start of study | Comparability of cohorts on the basis of the design or analysis | Assessment of outcome | Was follow up long enough for outcomes to occur | Adequacy of follow up of cohorts |  |
| Apea | n/a | n/a | * | * | ** | * | * | * | 7 |
| Aw | n/a | n/a | * | * | ** | * | * | * | 7 |
| Baker | n/a | n/a | * | * | ** | * | * | * | 7 |
| Brill | n/a | n/a | * | * |  | * | * | * | 5 |
| Cobos-Siles | n/a | n/a | * | * | ** | * | * | * | 7 |
| Crespo | n/a | n/a | * | * |  | * | * | * | 5 |
| De Smet | n/a | n/a | * | * | ** | * | * | * | 7 |
| Doglietto | n/a | n/a | * | * |  | * | * | * | 5 |
| Frost | n/a | n/a | * | * | ** | * | * | * | 7 |
| Hewitt | n/a | n/a | * | * | ** | * | * | * | 7 |
| Hoek | n/a | n/a | * | * |  | * | * | * | 5 |
| Knights | n/a | n/a | * | * |  | * | * | * | 5 |
| Miles | n/a | n/a | * | * | ** | * | * | * | 7 |
| Owen | n/a | n/a | * | * | ** | * | * | * | 7 |
| Rawle | n/a | n/a | * | * | ** | * | * | * | 7 |
| Thompson | n/a | n/a | * | * | ** | * | * | * | 7 |
| Conway | n/a | n/a | * | * |  | * |  | * | 4 |
| Steinmeyer | n/a | n/a | * | * |  | * | * | * | 5 |
| Kundi | n/a | n/a | * | * | ** | * | * | * | 7 |
| Hagg | n/a | n/a | * | * | ** | * | * | * | 7 |
| Marengoni | n/a | n/a | * | * | ** | * | * | * | 7 |
| Tehrani | n/a | n/a | * | * | ** | * | * | * | 7 |
| Mendes | n/a | n/a | * | * | * | * | * | * | 6 |
| Poco | n/a | n/a | * | * | * | * | * | * | 6 |
| Chinnadurai | n/a | n/a | * | * |  | * | * | * | 5 |
| Davis | n/a | n/a | * | * |  | * | * | * | 5 |

